# The Burden of Heart Failure in the US: Medical Costs and Health-Related Quality of Life

**DOI:** 10.1101/2025.10.31.25339280

**Authors:** Maria Alva, Sarahfaye Dolman, Slaven Sikirica, Paul Kolm, Katherine Andrade, Zugui Zhang, William S. Weintraub

## Abstract

**Background:** To quantify the economic burden and health-related quality of life (HRQoL) impact of heart failure (HF) in the U.S., stratified by age and diabetes status.

**Methods:** Using a micro-level, bottom-up prevalence-based approach from a payer perspective, we analyzed healthcare costs from the Optum Research Database (ORD). HF prevalence and HRQoL via EuroQol 5-Dimensions and Healthy Days instruments were estimated using National Health and Nutrition Examination Survey (NHANES).

**Results:** The average annual direct healthcare cost per HF patient was approximately $31,464 at baseline, rising to $45,893 in the first-year post-diagnosis before stabilizing around $37,500 annually. Inpatient care accounted for the largest cost share. Extrapolating at the national level, expenditures exceeded $227 billion at baseline and peaked at $332 billion in the year following a HF hospitalization. Individuals without diabetes had the highest quality-adjusted life years (QALYs) and lowest costs, while type 1 diabetes patients had the lowest QALYs and highest costs. Type 2 diabetes patients represented the largest subgroup with intermediate QALYs but substantial costs.

**Conclusions:** HF imposes a significant, sustained economic and quality-of-life burden, highlighting the need for targeted management strategies, especially for older adults and those with diabetes.

## Introduction

Heart failure (HF) is a chronic, progressive condition causing major functional limitations. Approximately 6.2 million adults in the US have HF^1^, and this number is projected to increase with population aging and increasing diabetes and hypertension^2,11^. HF is a leading cause of hospitalization and death. Roughly 870,000-900,000 new cases are diagnosed annually in the US^3^, and over 50% of patients die within five years of diagnosis^4,5^.

Although studies have documented the burden of HF-related hospitalizations and mortality, key limitations obscure true burden. Three recent systematic reviews^6–8^ highlight substantial variation in the cost components and estimates across U.S. studies, reflecting a lack of standardization. HF costs are typically divided into direct and indirect components. Direct costs include inpatient and outpatient services, procedures, and medications; indirect costs capture productivity losses, patient income loss, and unpaid caregiving. Most studies focus on direct medical costs and consistently identify hospitalizations—about two-thirds of total costs^7^—as the primary driver, largely due to the high burden among older adults. Aggregate-level analyses rarely account for cost differences across subgroups, such as those with diabetes, which worsens outcomes and increases expenses.

Population-based research is challenging because aggregate-level data that overlook cost differences among key subpopulations—such as those with coexisting diabetes, which worsens outcomes and raises costs—can mask meaningful variations.^9^ Also, HF often goes undiagnosed unless symptoms suddenly worsen. Chronic stable HF is more common but harder to detect, complicating prevalence estimation. Classification systems further contribute to variability; Rosamond, for example, found less than one-third overlap across diagnostic algorithms.^10^

Our study takes a targeted, policy-relevant approach by focusing exclusively on HF patients and examining the incremental burden of coexisting diabetes. Estimating HF’s causal impact on costs is difficult since it develops in older, sicker individuals with multiple comorbidities. By conditioning on HF and comparing those with and without diabetes, our analysis minimizes heterogeneity and clarifies the additional cost burden attributable to diabetes.

Using a large administrative database, we provide comprehensive estimates of the societal burden of HF in the U.S., including total and component-specific costs (inpatient, outpatient, and pharmaceutical) and health-related quality of life (HRQoL). Stratifying results by diabetes status and age highlights meaningful differences that can inform economic modeling, planning, and targeted interventions.

## Methods

### Data Sources

We used retrospective administrative claims data from the Optum Research Database (ORD) to examine the costs associated with comorbid diabetes (type 1 and 2) among patients with HF. The ORD is a database of administrative health claims for members of large commercial and Medicare Advantage health plans across all 50 states. ORD utilizes medical and pharmacy claims to derive patient-level enrollment information, health care costs, and resource utilization information. ORD administrative claims submitted for payment by providers and pharmacies are verified, adjudicated and de-identified prior to inclusion. Mortality data were also included from a variety of sources, including the Social Security Administration’s Death File. The study was IRB-exempt and did not require informed consent.

Table S1 shows the complete list of ICD-10 codes used to identify HF^11^ among adults (18+) hospitalized between January 1, 2016, and December 30, 2023. The index date was defined as discharge from the first hospitalization with HF as the primary diagnosis. The start year coincided with the ICD-9 to ICD-10 transition to ensure coding consistency. We limited the sample to acute care hospitalizations to standardize disease severity and clinical context, and to reduce misclassification due to incidental or secondary HF diagnoses.

We required patients to have at least 12 months of continuous medical and pharmacy enrollment prior to the index date to allow for robust baseline characterization. Given the high death rate in this population, we required only one day of follow-up continuous medical and pharmacy enrollment following the index date. However, for the yearly utilization and cost tables, we required continuous medical and pharmacy enrollment for the given number of years or less if due to death following the index date.

To reduce heterogeneity, we excluded hospitalizations lasting >30 days, admissions with major concurrent events (e.g., stroke, dialysis), and patients with prior SGLT2 inhibitor use due to their impact on HF outcomes. Patients were followed for up to five years, death, end of study period (December 31, 2023), or disenrollment.

To estimate HF prevalence and HRQoL, we used National Health and Nutrition Examination Survey (NHANES) data, identifying HF through questionnaire responses. CDC Healthy Days data were mapped to EuroQol 5-Dimensions (EQ-5D) utility scores to compare HRQoL between individuals with and without HF, following prior methods^12^. American Community Survey (ACS) population counts by age were combined with NHANES HF prevalence to estimate the number of adults with HF nationally, allowing us to scale patient-level costs to the population level.

### Study design and statistical analyses

We used a prevalence-based, bottom-up approach to estimate HF costs from the payer perspective, focusing on direct medical costs in 2023 dollars^13,14^. We adjust for inflation using medical care component of the Consumer Price Index, ensuring comparability over time by expressing values in constant dollars^15^.

Multiplying ACS counts by NHANES prevalence yields the estimated number of people with HF. We then link these population estimates with average per-patient healthcare cost data from the Optum claims database, which captures real-world medical expenditures for individuals with HF. Multiplying the HF population by the per-patient costs produces total annual national costs stratified by age, cost type, with and without comorbid diabetes, and year to reflect expected cost trajectories.

We estimate the immediate (acute) costs incurred in the year of HF onset and the longer-term costs in subsequent years (years 2–5), recognizing that the economic burden of HF evolves over time. Patients’ enrollment in the health plan is divided into 12-month periods following their index date. Cost measurement is conducted only for intervals in which the patient maintained continuous enrollment for the full 12 months, unless the interval is truncated due to death. Patients are observed until they disenroll from the health plan or reach the end of the study period. Costs are measured directly and include both health plan payments and patient out-of-pocket amounts. Costs are HF-related if there was a diagnosis code for HF in any position or if there was a HF medication. Patients who are censored due to disenrollment or the end of the study period are **excluded** from the corresponding follow-up year’s cost analysis. Patients who die during a follow-up year are **included** in that year’s cost analysis, as complete cost data are available for the time they were enrolled, even if it is less than a full year.

Annual average costs following diagnosis are estimated by multiplying the Kaplan–Meier (KM) survival probability at the start of the interval by the estimated average costs during that interval, conditional on survival. The total average cost over the five-year period is obtained by summing these interval-specific estimates^16,17^. To estimate the total population-level burden of HF, for each period, we first calculate the average cost per patient per year by summing the product of cost and survival probability across periods, and dividing by the number of patients at the start of the study. This accounts for censoring and differential survival. Next, we use published prevalence estimates of HF obtained from NHAHES and apply these to U.S. Census population data to estimate the total number of individuals living with HF in the population. Finally, we multiply the average total cost per patient by the estimated number of people with HF to derive the total direct medical cost burden of the disease at the national level.

To estimate mortality and quality-adjusted life expectancy for patients with HF, we used the KM product-limit estimator to calculate survival probabilities which accounts for right-censoring, ensuring that death rates are not underestimated due to loss to follow-up or study period truncation^18^. We then calculated the overall probability of survival over five years as the product of period-specific survival probabilities, i.e., *P*_1_ ×*P*_2_ ×*P*_3_ ×*P*_4_ ×*P*_5_ where *P*_*i*_ is the probability of serving at time *i*.

To compute quality-adjusted life-years (QALYs), we multiplied survival probabilities for each period by corresponding estimates of quality of life (*QoL*), incorporating a decrement over time to reflect the progressive nature of HF as follows:

Period 1=*P*_1_× *QoL*_1_
Period 2==*P*_1_×*P*_2_ × *QoL*_2_
Period 3==*P*_1_×*P*_2_ × *P*_3_ × *QoL*_3_

We assume a baseline QoL of 0.85, consistent with mild-to-moderate impairment. For each additional year, we applied a decrement of 0.107, drawn from published estimates of utility loss associated with HF progression^12^. These decrements are based on mappings from Healthy Days measures in the NHANES survey to the EQ-5D utility scale, following the method proposed by Jia and Lubetkin^19^ -- a regression-based mapping which provides preference-based estimates for EQ-5D scores from self-reported physically and mentally unhealthy days. Using this approach, we derived utility weights for each follow-up period and summed the period-specific products of survival and QoL to obtain total QALYs. This framework integrates both longevity and HRQoL to support burden-of-disease and cost-effectiveness analyses.

## Results

The CONSORT table (Figure S1), outlines the sample construction process. We began with approximately 38 million commercial and Medicare Advantage health plan members over the age of 18 with at least one day of medical and pharmacy enrollment during the identification period (January 01, 2016, to December 30, 2023). Nearly 500,000 or 1.32% experienced a hospitalization with HF as the primary diagnosis. The discharge date was set as the index date. After applying the inclusion/ exclusion criteria, the final analytic sample included 258,096 patients, of whom nearly half (49.7%) had diabetes, with 9.3% having type 1 diabetes and 90.7% having type 2 diabetes. 200,101 patients were observed for at least one full year, and 19,248 for the full five-year period.

Table S2 summarizes the demographic, clinical, and treatment characteristics of the Optum cohort. The overall mean age is 75 years, ranging from 71.9 years in the type 1 diabetes group to 75.5 years among those without diabetes. Comorbidities such as coronary artery disease, peripheral artery disease, and chronic kidney disease were more common in both diabetes groups—especially type 1—than in those without diabetes. Cardiovascular medications (RAAS inhibitors, beta blockers, diuretics) were widely prescribed across all groups. The Charlson comorbidity score, indicating disease burden, was highest for type 1 diabetes (4.31), followed by type 2 (3.75) and no diabetes (2.61).

Table S3 shows baseline total direct medical costs per person (mean, median, and standard deviation (SD)) in the year preceding the first HF hospitalization. Mean total healthcare cost for individuals with HF in the 12-months prior to the index hospitalization is $31,463.56 (median: $16,299.98). Patients aged 18–44 years have an average annual cost of $41,850.28 (median: $9,597.38), those aged 45–64 years have the highest annual cost equal to $43,306.62 (median: $19,566.54), and those aged 65–74 have on average costs equal to $31,178.45 (median: $17,257.54). Among patients aged 75 and older, the average annual cost is lowest, at $27,093.92 (median: $15,416.07). The higher mean but lower median within younger age groups suggests substantial cost variation. When considering HF-related costs specifically, the overall mean is $7,513.34 (median: $386.24). These costs also vary by age group, with the highest mean HF-related expenses observed among individuals aged 45–64 years ($10,319.30) and 18–44 years ($10,019.80), and the lowest among those aged 75 and older ($6,561.87). Inpatient stays represent the largest cost component across all age groups at both baseline and all subsequent years. At baseline, IP costs represent 31.5% of total costs for people with HF. Ambulatory costs follow closely at 29.1%. Pharmacy costs represent 23.0% of total costs. Inpatient stays are even a larger driver of HF-specific costs (63.5%), whereas the opposite is true for outpatient care (17.3%) and pharmacy costs (3.9%) specific to HF only.

Other medical costs have the highest variation (SD/Mean = 8.89) followed by outpatient visits (4.66) and ambulatory care (4.21). Office visits (2.76) and pharmacy costs (2.77) have the lowest relative variation, suggesting that these are more predictable expenses.

Figure 1 shows the total average direct healthcare costs associated with HF at baseline and up to 5 years post-hospitalization, along with 95% confidence intervals, and where the sample in each follow-up year includes those with the given length of continuous enrollment or less if due to death. In the year preceding the HF hospitalization (Year 0), HF-related costs are approximately $10,000, while all-cause costs are around $30,000. Costs increase sharply in the year that starts with the indexed hospitalization (Year 1), with all-cause costs peaking above $45,000 and HF-related costs approaching $28,000. After Year 1, both cost categories decline but remain elevated compared to Year 0. By Years 2 through 5, all-cause costs stabilize between $35,000 and $38,000 annually, while HF-related costs stabilize around $18,000 to $20,000 per year. This pattern highlights the substantial economic burden following HF diagnosis, with the first-year post-diagnosis associated with the highest costs, particularly driven by all-cause healthcare utilization. Approximately 23.9% (7,513.34/31,463.56) of the total healthcare costs for people with HF are related to HF alone at baseline, underscoring that much of the spending stems from coexisting conditions.

**Figure 1:**
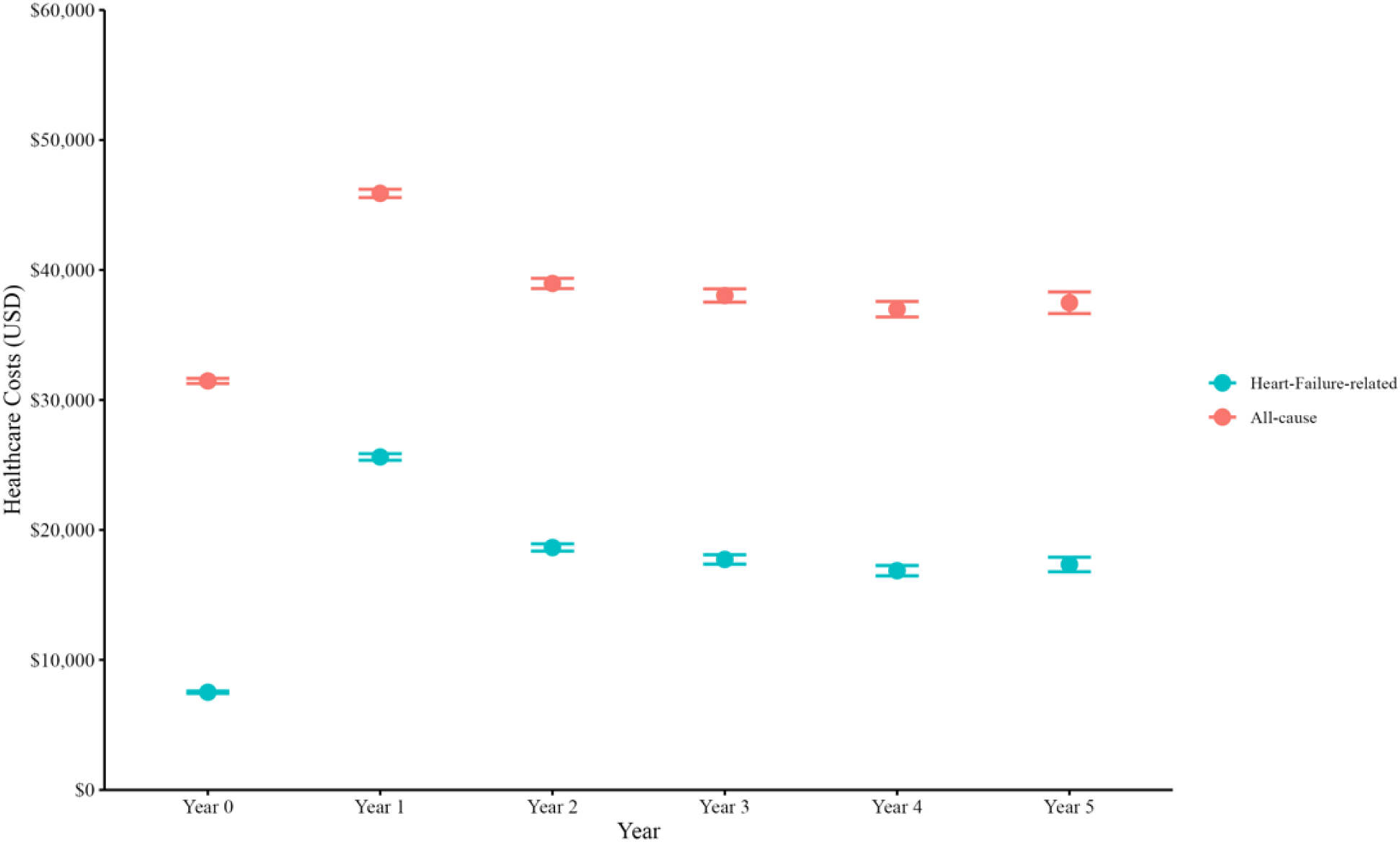
Annual Heart-Failure-Related and All-Cause Healthcare Costs in the Five Years Following Heart Failure Diagnosis *Note: the patient sample in each year during the follow-up period (years 1-5) includes patients with the given length of continuous enrollment, or less if due to death. Patients who die are not included in subsequent yearly estimates*.

Table 1 shows that outpatient care costs remain stable, increasing slightly from $12,286.78 at baseline to $13,020.98 in year 5, and consistently accounting for about 34–39% of total annual healthcare costs. Pharmacy costs follow a similar trend, rising from $7,254.15 to $8,432.72, accounting for 22.5-23.1% share. In contrast, emergency and acute care costs show greater fluctuation, peaking in year 1 at $23,455.22 (51.1%), before declining to $16,026.93 (42.8%) in year 5. These shifts reflect the intense resource demands immediately after HF hospitalization, followed by a gradual redistribution of costs toward outpatient and pharmacy care. Patients with type 1 diabetes had the highest total costs, followed by type 2, while those without diabetes had the lowest. At baseline, pharmacy costs were 117% higher for type 1 and 48% higher for type 2 compared with non-diabetic patients. By year 5, these remained 80% and 54% higher, reflecting the ongoing burden of medication management, especially for insulin users.

**Table 1:**
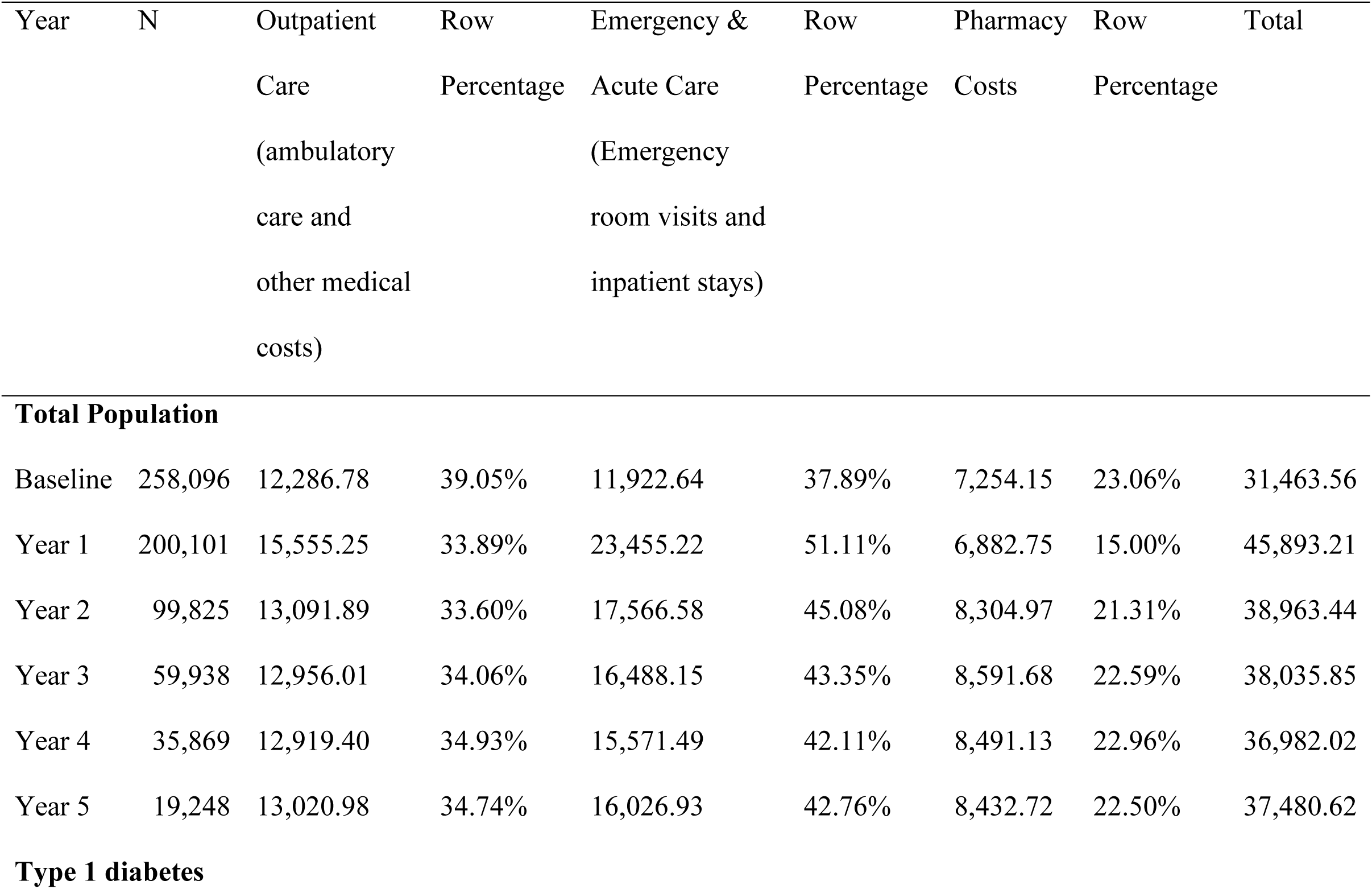
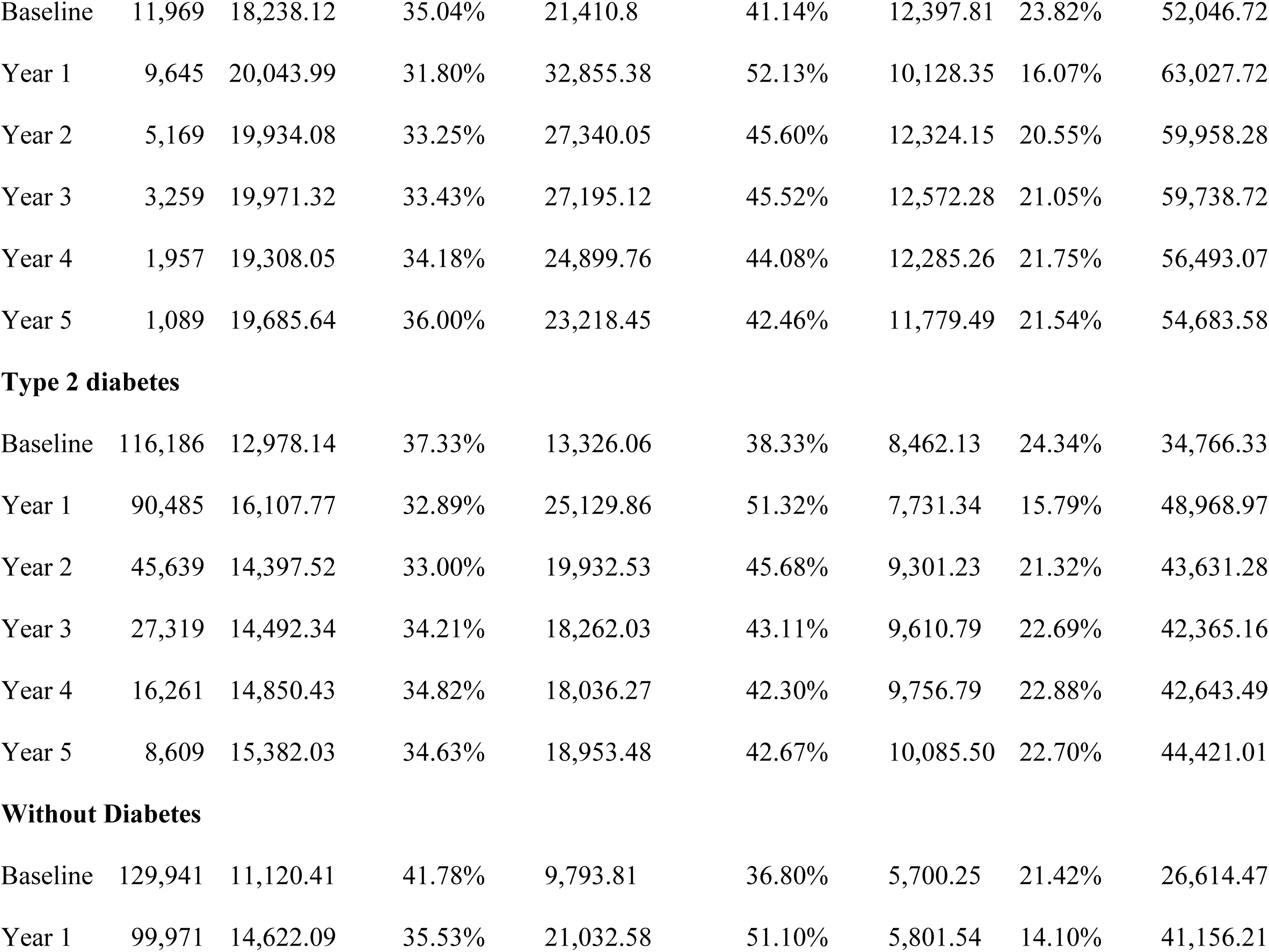
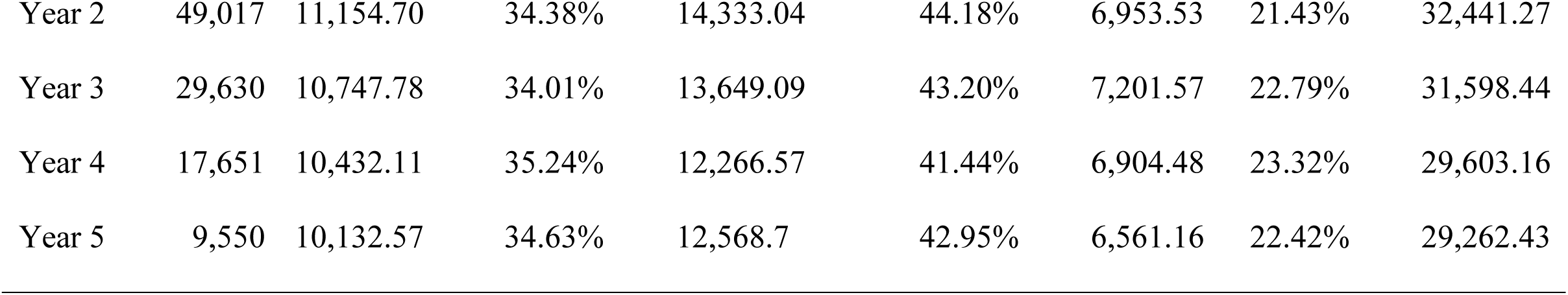
Annual burden per person in the first year and in subsequent years by utilization type and diabetes status.

| Year | N | Outpatient<br>Care<br>(ambulatory<br>care and<br>other medical<br>costs) | Row<br>Percentage | Emergency &<br>Acute Care<br>(Emergency<br>room visits and<br>inpatient stays) | Row<br>Percentage | Pharmacy<br>Costs | Row<br>Percentage | Total |
| --- | --- | --- | --- | --- | --- | --- | --- | --- |
| Total Population |  |  |  |  |  |  |  |  |
| Baseline | 258,096 | 12,286.78 | 39.05% | 11,922.64 | 37.89% | 7,254.15 | 23.06% | 31,463.56 |
| Year 1 | 200,101 | 15,555.25 | 33.89% | 23,455.22 | 51.11% | 6,882.75 | 15.00% | 45,893.21 |
| Year 2 | 99,825 | 13,091.89 | 33.60% | 17,566.58 | 45.08% | 8,304.97 | 21.31% | 38,963.44 |
| Year 3 | 59,938 | 12,956.01 | 34.06% | 16,488.15 | 43.35% | 8,591.68 | 22.59% | 38,035.85 |
| Year 4 | 35,869 | 12,919.40 | 34.93% | 15,571.49 | 42.11% | 8,491.13 | 22.96% | 36,982.02 |
| Year 5 | 19,248 | 13,020.98 | 34.74% | 16,026.93 | 42.76% | 8,432.72 | 22.50% | 37,480.62 |
| Type 1 diabetes |  |  |  |  |  |  |  |  |
| Baseline | 11,969 | 18,238.12 | 35.04% | 21,410.8 | 41.14% | 12,397.81 | 23.82% | 52,046.72 |
| Year 1 | 9,645 | 20,043.99 | 31.80% | 32,855.38 | 52.13% | 10,128.35 | 16.07% | 63,027.72 |
| Year 2 | 5,169 | 19,934.08 | 33.25% | 27,340.05 | 45.60% | 12,324.15 | 20.55% | 59,958.28 |
| Year 3 | 3,259 | 19,971.32 | 33.43% | 27,195.12 | 45.52% | 12,572.28 | 21.05% | 59,738.72 |
| Year 4 | 1,957 | 19,308.05 | 34.18% | 24,899.76 | 44.08% | 12,285.26 | 21.75% | 56,493.07 |
| Year 5 | 1,089 | 19,685.64 | 36.00% | 23,218.45 | 42.46% | 11,779.49 | 21.54% | 54,683.58 |
| **Type 2 diabetes** |  |  |  |  |  |  |  |  |
| Baseline | 116,186 | 12,978.14 | 37.33% | 13,326.06 | 38.33% | 8,462.13 | 24.34% | 34,766.33 |
| Year 1 | 90,485 | 16,107.77 | 32.89% | 25,129.86 | 51.32% | 7,731.34 | 15.79% | 48,968.97 |
| Year 2 | 45,639 | 14,397.52 | 33.00% | 19,932.53 | 45.68% | 9,301.23 | 21.32% | 43,631.28 |
| Year 3 | 27,319 | 14,492.34 | 34.21% | 18,262.03 | 43.11% | 9,610.79 | 22.69% | 42,365.16 |
| Year 4 | 16,261 | 14,850.43 | 34.82% | 18,036.27 | 42.30% | 9,756.79 | 22.88% | 42,643.49 |
| Year 5 | 8,609 | 15,382.03 | 34.63% | 18,953.48 | 42.67% | 10,085.50 | 22.70% | 44,421.01 |
| **Without Diabetes** |  |  |  |  |  |  |  |  |
| Baseline | 129,941 | 11,120.41 | 41.78% | 9,793.81 | 36.80% | 5,700.25 | 21.42% | 26,614.47 |
| Year 1 | 99,971 | 14,622.09 | 35.53% | 21,032.58 | 51.10% | 5,801.54 | 14.10% | 41,156.21 |
| Year 2 | 49,017 | 11,154.70 | 34.38% | 14,333.04 | 44.18% | 6,953.53 | 21.43% | 32,441.27 |
| Year 3 | 29,630 | 10,747.78 | 34.01% | 13,649.09 | 43.20% | 7,201.57 | 22.79% | 31,598.44 |
| Year 4 | 17,651 | 10,432.11 | 35.24% | 12,266.57 | 41.44% | 6,904.48 | 23.32% | 29,603.16 |
| Year 5 | 9,550 | 10,132.57 | 34.63% | 12,568.7 | 42.95% | 6,561.16 | 22.42% | 29,262.43 |

Outpatient care costs for individuals with type 1 diabetes were 64% higher than those without diabetes at baseline, and 94% higher by year 5. For type 2 diabetes, the increases were 17% and 52%, reflecting greater reliance on ambulatory and specialist care.

Emergency and acute care costs also remained higher for individuals with diabetes. At baseline, those with type 1 diabetes incurred 119% higher costs than those without diabetes; for type 2, the gap was 36%. These differences persisted at year 5, with emergency and acute care costs 85% higher for type 1 and 51% higher for type 2, indicating a sustained burden of complications and hospital-based care.

At the population level (Table 2), individuals aged 45–64 years account for the highest total medical direct costs, from $77.07 billion at baseline to $131.60 billion in year 1, with costs remaining above $96 billion annually through year (Table 2). Younger adults aged 18–44 years, despite a low HF prevalence of 0.50%, also incur substantial costs, particularly in the first year of their indexed hospitalization, with total costs rising from $25.23 billion at baseline to $55.77 billion in year 1. In contrast, older adults aged 65–74 and 75+ years—who have the highest HF prevalence (6.57% and 11.04%, respectively)—peak at $117.44 billion and $98.24 billion in year 1, respectively. A similar age pattern emerges for HF-related costs only (Table S3). Although adults aged 75 and older have the highest HF prevalence (11.0%), their HF-related costs are lower than those of middle-aged adults. At the population level, total HF-related expenditures rise sharply from $54.3 billion at baseline to $185.3 billion in year 1, then decline and stabilize around $125 billion by year 5. These trends underscore the substantial economic burden of HF across all ages, with resource use peaking in the first year after diagnosis and among adults aged 45–64.

**Table 2:** Trends in Total and Heart Failure-Related Costs by Age Group Over Five Years.

| Characteristic | Total | Prevalence | Baseline | Year 1 | Year 2 | Year 3 | Year 4 | Year 5 |
| --- | --- | --- | --- | --- | --- | --- | --- | --- |
| | Population<br>(ACS*) | (NHANES<sup>†</sup>, %) | (\$ billions) | (\$ billions) | (\$ billions) | (\$ billions) | (\$ billions) | (\$ billions) |
| Total Healthcare Costs for People with Heart Failure |  |  |  |  |  |  |  |  |
| 18 - 44 | 120,574,085 | 0.50% | \$25.23 | \$55.77 | \$37.06 | \$40.05 | \$34.26 | \$38.48 |
| 45 - 64 | 82,395,715 | 2.16% | \$77.07 | \$131.60 | \$103.60 | \$98.27 | \$93.91 | \$96.64 |
| 65 - 74 | 34,824,818 | 6.57% | \$77.05 | \$117.44 | \$95.53 | \$93.22 | \$90.84 | \$90.57 |
| 75+ | 24,482,238 | 11.04% | \$73.23 | \$98.24 | \$85.45 | \$83.30 | \$81.21 | \$80.88 |
| All | 231,791,117 | 3.12% | \$227.54 | \$331.89 | \$281.78 | \$275.07 | \$267.45 | \$271.06 |
| Heart Failure Related Costs for People with Heart Failure |  |  |  |  |  |  |  |  |
| 18 - 44 | 120,574,085 | 0.50% | \$6.04 | \$29.91 | \$13.99 | \$17.90 | \$14.40 | \$20.82 |
| 45 - 64 | 82,395,715 | 2.16% | \$18.37 | \$71.49 | \$49.34 | \$44.87 | \$40.27 | \$40.05 |
| 65 - 74 | 34,824,818 | 6.57% | \$18.00 | \$64.33 | \$44.12 | \$42.73 | \$41.28 | \$41.69 |
| 75+ | 24,482,238 | 11.04% | \$17.74 | \$56.26 | \$42.47 | \$39.84 | \$38.44 | \$39.42 |
| All | 231,791,117 | 3.12% | \$54.34 | \$185.25 | \$134.87 | \$128.23 | \$121.97 | \$125.42 |

\*American Community Survey, <sup>†</sup>National Health and Nutrition Examination Survey

QALYs decrease from 0.61 in year 1 (71.52% survival time 0.85 assumed utility) to 0.29 by year 5 (assuming an estimate 0.107 disutility based on NHANES), indicating that both survival and HRQoL deteriorate significantly over time. Individuals with type 1 diabetes consistently exhibit the lowest QALY estimates. By year 5, their QALYs decline to 0.25, compared to 0.30 among individuals without diabetes. Figure 2 illustrates the decline in QALYs over a five-year period following a HF hospitalization, comparing individuals with type 1 diabetes and individuals without diabetes. Results for individuals with type 2 diabetes are not shown, as their QALY estimates are virtually identical to those for individuals with type 1 diabetes, making the curves indistinguishable on this scale. Here we assumed the same HrQoL (disutility) for individuals with and without diabetes, making these estimates highly conservative. In reality, people with diabetes—particularly those with complications—are likely to experience greater disutility and lower QALYs than individuals without diabetes. Therefore, the true difference in QALYs between individuals with and without diabetes is likely larger than shown here. Across both groups, there is a marked decline in QALYs over time following a HF hospitalization, with the steepest reduction observed in year 1. This is not surprising because the largest decrement in survival (28.5%) happens within the first year following a hospitalization. Subsequent marginal increments in cumulative probability of death tend to be smaller, reflecting that the highest-risk individuals tend to die earlier, and the remaining population is somewhat "healthier" or more resilient (a form of survivor bias as shown in Table S4).

**Figure 2:**
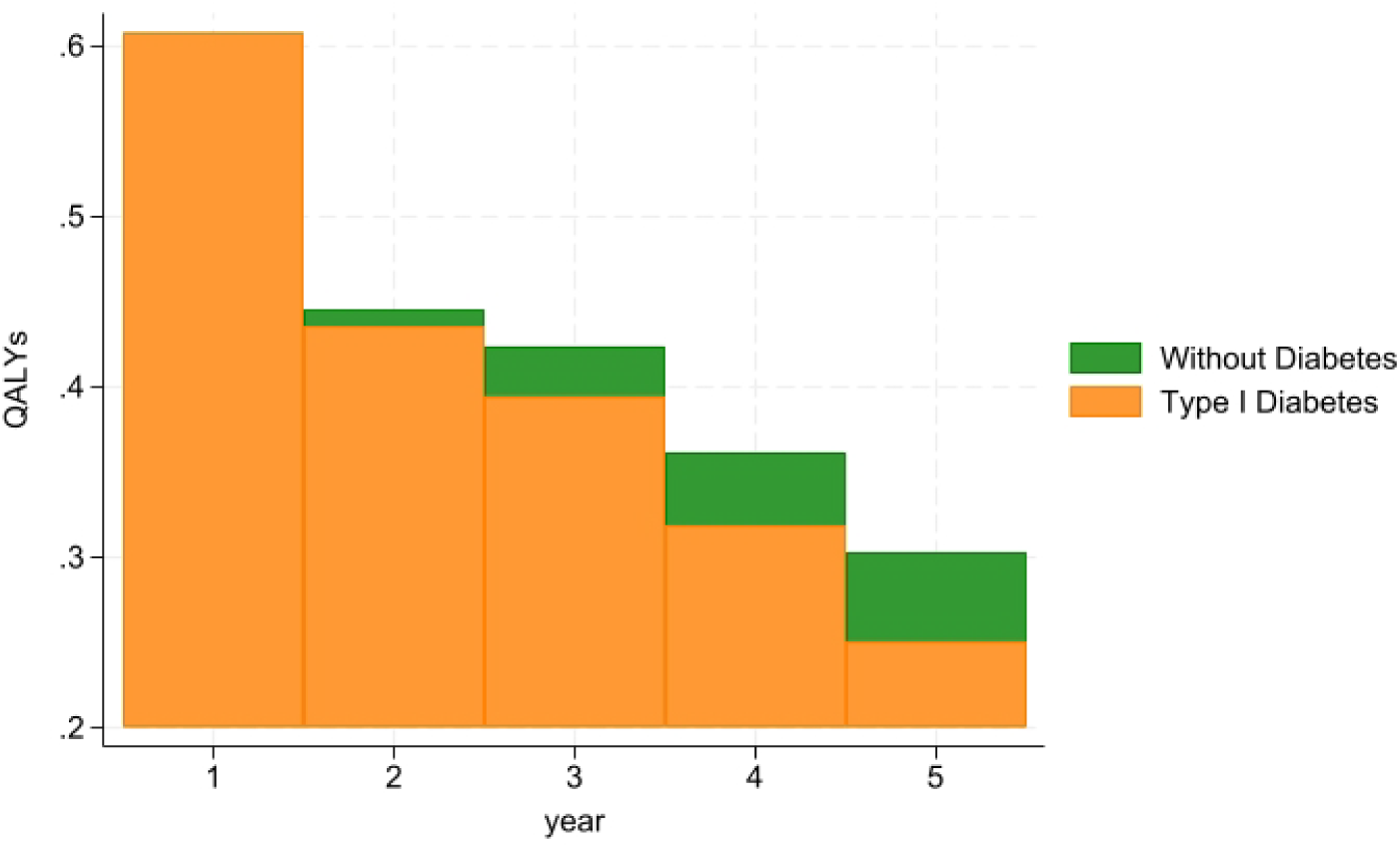
QALYs Over Five Years for People with and without Diabetes Following HF Hospitalization

## Discussion

This study provides a comprehensive assessment of the economic and HRQoL burden associated with HF within a large, insured U.S. population. Our findings reveal significant direct healthcare costs and substantial declines in QALYs following HF hospitalization, with notable differences by age and diabetes status. These results underscore the persistent and multifaceted burden of HF on both the healthcare system and affected individuals.

Healthcare costs peaked in the year after hospitalization, driven largely by emergency and acute care, before stabilizing above baseline. Younger adults (18–64) incurred higher average costs than older adults, reflecting more aggressive, resource-intensive treatments such as defibrillators, assist devices, and advanced drugs. In contrast, frailty and limited life expectancy among those aged 75+ likely reduce intervention intensity and spending.

Across all periods, individuals with type 1 diabetes incurred the highest total healthcare costs, with consistently elevated spending across outpatient care, emergency and acute care, and pharmacy categories. Pharmacy costs for this group were nearly double those of individuals without diabetes, reflecting the complexity of disease management, including lifelong insulin therapy. Type 2 diabetes was also associated with higher healthcare costs, though the gap was less pronounced. Importantly, these cost differentials were accompanied by lower QALY estimates for individuals with diabetes, particularly those with type 1 diabetes, underscoring the compounded economic and health burden of managing coexisting HF and diabetes. Notably, our application of equal disutility assumptions for individuals with and without diabetes likely results in conservative QALY estimates. In reality, diabetes-specific complications and comorbidities likely contribute to greater reductions in HRQoL, suggesting that our results underestimate the true burden faced by this population.

Our findings are within the thresholds of prior U.S.-based studies. As shown in Table S5, existing U.S. cost-of-illness estimates for HF vary considerably, with differences in total healthcare costs reaching nearly $150,000 and hospitalization costs differing by over $110,000. This wide variation likely reflects differences in study design, perspective (e.g., payer vs. societal), resource utilization patterns, payer type and underlying population characteristics. Specifically, differences in healthcare costs observed between commercially insured individuals and those covered by Medicare (65+) may reflect Medicare’s stronger negotiating power, resulting in lower reimbursement rates and, consequently, reduced expenditures. This points to the potential value of policy approaches that leverage purchasing power to contain costs across payers. Despite these differences, the consistent observation across studies of high costs in the first year following HF hospitalization and sustained elevated costs thereafter supports the robustness of our findings.

Strengths of this study include the large, contemporary, and nationally representative insured population, extended five-year follow-up, and detailed cost stratification by age and diabetes status. Some limitations warrant consideration. First, the lack of a comparison group without HF limits our ability to isolate the incremental cost and health burden attributable solely to HF. Second, while our administrative claims data enable precise cost estimation, they lack clinical detail on HF severity, treatment adherence, and functional status, all of which may influence both costs and outcomes. Lastly, as noted, our QALY estimates are based on conservative assumptions regarding diabetes-related disutility, which may potentially underestimate the true quality of life burden in this high-risk group.

## Conclusions

At the national level, HF drives healthcare spending exceeding $227 billion annually, rising to $332 billion in the first year after diagnosis. Although costs decline in subsequent years, they remain elevated, reflecting the sustained demands of HF management. Adults aged 45–64 account for a disproportionate share, emphasizing the need for age-targeted strategies to reduce hospitalizations and improve outcomes. Individuals with type 1 diabetes experience lower QALYs than those without diabetes, underscoring the added burden of comorbidity.

## Data Availability

This study used de-identified administrative claims data from the Optum Research Database (ORD) and publicly available datasets from the National Health and Nutrition Examination Survey (NHANES) and the American Community Survey (ACS). The ORD data were accessed under a data use agreement and are not publicly available due to licensing restrictions. NHANES and ACS data are publicly accessible through the U.S. Centers for Disease Control and Prevention and the U.S. Census Bureau, respectively.

https://www.census.gov/programs-surveys/acs

https://wwwn.cdc.gov/nchs/nhanes/

https://www.optum.com/business/resources.html

## Supplemental Material

Tables S1-S5

Figure S1

## Abbreviations

ACS: American Community Survey
EQ-5D: EuroQol 5-Dimensions
HF: Heart failure
HRQoL: Health-related quality of life
KM: Kaplan–Meier
NHANES: National Health and Nutrition Examination Survey
ORD: Optum Research Database
QALY: Quality-adjusted life year

